# SARS-CoV-2 antibody responses determine disease severity in COVID-19 infected individuals

**DOI:** 10.1101/2020.07.27.20162321

**Authors:** Cecilie Bo Hansen, Ida Jarlhelt, Laura Pérez-Alós, Lone Hummelshøj Landsy, Mette Loftager, Anne Rosbjerg, Charlotte Helgstrand, Jais Rose Bjelke, Thomas Egebjerg, Joseph G. Jardine, Charlotte Sværke Jørgensen, Kasper Iversen, Rafael Bayarri-Olmos, Peter Garred, Mikkel-Ole Skjoedt

## Abstract

Globally, the COVID-19 pandemic has had extreme consequences for the healthcare system and calls for diagnostic tools to monitor and understand the transmission, pathogenesis and epidemiology, as well as to evaluate future vaccination strategies. Here we have developed novel flexible ELISA-based assays for specific detection of SARS-CoV-2 antibodies against the receptor-binding domain (RBD): An antigen sandwich-ELISA relevant for large population screening and three isotype-specific assays for in-depth diagnostics. Their performance was evaluated in a cohort of 350 convalescent participants with previous COVID-19 infection, ranging from asymptomatic to critical cases. We mapped the antibody responses to different areas on protein N and S and showed that the IgM, A and G antibody responses against RBD are significantly correlated to the disease severity. These assays—and the data generated from them—are highly relevant for diagnostics and prognostics and contribute to the understanding of long-term COVID-19 immunity.

## Introduction

Coronaviruses (CoVs) are zoonotic pathogens primarily targeting the human respiratory system (1). While most human CoV infections are mild, three coronaviruses have appeared in the past two decades that cause deadly pneumonia. Severe acute respiratory syndrome coronavirus (SARS-CoV-1), first observed in China in 2002, spread rapidly to 26 countries infecting more than 8,000 people and causing 774 deaths before being contained in 2003 (2). Barely a decade later in 2012, Middle East respiratory syndrome coronavirus (MERS-CoV) was identified on the Arabian peninsula and has caused more than 2,500 cases and 866 deaths (3–5). At the end of 2019, a novel SARS-CoV strain (SARS-CoV-2) emerged in Wuhan (China) and has been spreading at an unprecedented speed around the world ever since. The disease, named Coronavirus Disease 2019 (COVID-19), accounts for more than 15 million confirmed cases and 630,000 related deaths at the time of manuscript preparation (6). The clinical features of the COVID-19 infection are diverse, ranging from asymptomatic carriers of the infection to acute respiratory distress syndrome and multiple organ dysfunction (7). SARS-CoV-2 can infect people of all ages; however, elderly and people suffering from co-morbidities such as diabetes, cardiovascular disease, chronic respiratory disease or cancer are more inclined to suffer from a severe disease progression (8).

SARS-CoV-2 is an enveloped RNA virus with a diameter of 65–125 nm to accommodate one of the largest genomes of all known RNA viruses (29.8–29.9 kb) (9). One-third of its genome encodes four structural proteins: spike (S), membrane (M), envelope (E) and nucleocapsid (N) proteins. The protein S that extends as homotrimers on the outer viral membrane binds to the host angiotensin-converting enzyme 2 (ACE2) receptor and allows the virus to enter and infect the host target cell (10). Host proteases process the protein S into the S1 and S2 subunits: S1 is responsible for receptor recognition and is comprised of an N-terminal and a C-terminal domain, the later containing the receptor-binding domain (RBD) (11); while the S2 mediates the fusion of the viral envelope with the membrane of the host cell. The protein S of SARS-CoV-2 shares 76% homology at the amino acid level with the protein S of SARS-CoV-1 and while both interact with the ACE2 receptor, SARS-CoV-2 does so with a 10–20 times higher affinity (12), which may explain the higher transmission rate of COVID-19.

Precise diagnosis of COVID-19 with RT-PCR detection of SARS-CoV-2 nucleic acids remains crucial to identify symptomatic carriers to secure correct treatment and as a tool in quarantine strategies to limit the infection rate in asymptomatic carriers. Serological detection of specific SARS-CoV-2 antibodies is a useful tool to identify convalescent individuals that have developed immunity and thereby might be protected against reinfection, although this issue is still not resolved (13). Moreover, serological testing is necessary to understand the transmission, pathogenesis and epidemiology of SARS-CoV-2, providing critical data to inform public health authorities for controlling the spread of COVID-19 and eventually re-opening societies. Another question that has arisen is whether an overwhelming antibody response may even aggravate COVID-19 infection in patients (14).

Due to urgency and demand, many serological tests have been developed rapidly and made commercially available with only limited validation on clinical samples (15). We have developed a flexible ELISA-based platform for rapid and specific detection of SARS-CoV-2 antibodies. The platform includes an indirect RBD sandwich ELISA (S-ELISA) for pan immunoglobulin (Ig) detection suitable for large scale antibody surveillance and direct ELISAs for in-depth analyses of the IgM, IgA and IgG isotype antibody responses towards RBD and protein N. Moreover, we set out to characterize the antibody response levels in relation to symptom characteristics and disease severity, to elucidate the immunological response in COVID-19 convalescent individuals.

## Results

### 1. Development of specific ELISA-based assays for the detection of antibodies against SARS-CoV-2

The assays were optimized with a specific focus on diminishing nonspecific binding and the final setup was chosen based on the optimal intensity (OD) of the absorbance signal and signal-to-noise (S/N) ratio between the positive sample and the negative quality control. The developed ELISA-based assays proved to be suitable for automatization and were used in high-throughput setups with both 96-well and 384-well formats, correlating significantly (Supplementary Figure 1A–D). No differences were observed when serum or plasma was employed for the detection of human antibodies against SARS-CoV-2 (Supplementary Figure 1E–G).

**Figure 1.**
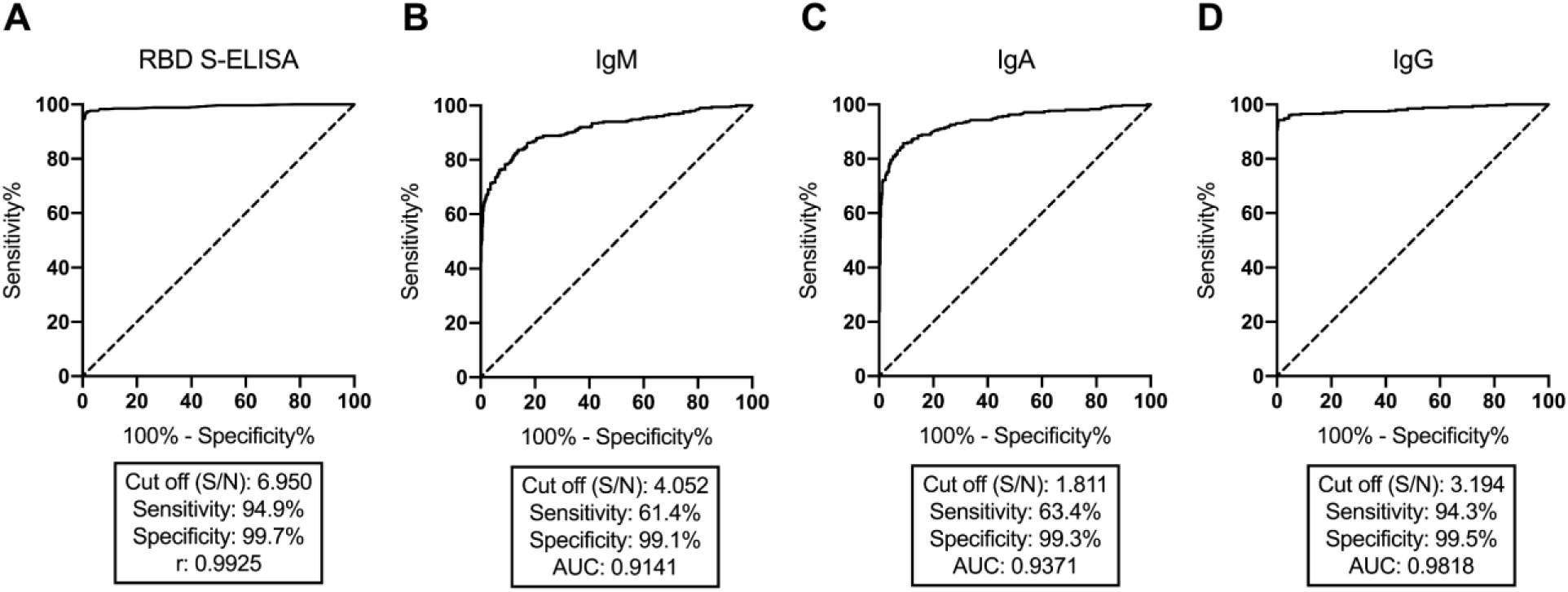
Assay validation. ROC curves analysis representing the performance of the RBD S-ELISA setup detecting total Ig (A) and the direct ELISA setup detecting IgM (B), IgA (C) and IgG (D) using RBD as antigen. A total of 350 positive samples and 580 negative controls were subjected to antibody detection using the 384-well format. AUC: Area under the curve.

### 2. Assay validation

A total of 350 convalescence plasma samples from previously infected individuals with SARS-CoV-2 (positive samples) and 580 plasma samples from healthy individuals collected before the pandemic outbreak (negative controls) were subjected to antibody measurement in the RBD S-ELISA (Figure 1A) and in the direct ELISA setups (Figure 1B– D). S/N ratio between the OD of a positive sample and the OD of the negative quality control and receiver operating characteristic (ROC) were assessed to calculate the best fit cut off to estimate the performance of the assay. The RBD S-ELISA performed with a 94.9% sensitivity and 99.7% specificity (Figure 1A). The sensitivities and specificities of the direct ELISAs were 61.4%, 99.1% for IgM (Figure 1B), 63.4%, 99.3% for IgA (Figure 1C) and 94.3%, 99.5% for IgG (Figure 1D), respectively. The intra- and inter-assay variation were found to be acceptable (< 10%) for all four assays (Table 1). The limit of detection was determined by interpolating the cut off OD value and converting it into antibody concentrations. The resulting values indicate an estimated 10-fold higher sensitivity of the direct ELISA setup (0.406 ng/ml) compared to the S-ELISA (4.046 ng/ml) (Table 1).

**Table 1.** Intra- and inter-assay variation and limit of detection for the S-ELISA setup and the direct ELISA setup.

| ASSAY | n | SD | CV (%) | ng/ml |
| --- | --- | --- | --- | --- |
| **Intra-assay variation** |  |  |  |  |
| **RBD S-ELISA** | 40 | 0.045 | 5.9 | - |
| **Direct ELISA** |  |  |  |  |
| **IgM** | 40 | 0.004 | 5.4 | - |
| **IgA** | 40 | 0.006 | 5.8 | - |
| **IgG** | 40 | 0.020 | 6.3 | - |
| **Intra-assay variation** |  |  |  |  |
| **RBD S-ELISA** | 24 | 0.057 | 7.6 | - |
| **Direct ELISA** |  |  |  |  |
| **IgM** | 24 | 0.065 | 8.6 | - |
| **IgA** | 24 | 0.126 | 7.5 | - |
| **IgG** | 24 | 0.154 | 8.4 | - |
| **Limit of detection** |  |  |  |  |
| **RBD S-ELISA** | - | - | - | 4.046 |
| **Direct ELISA** | - | - | - | 0.406 |

CV: Coefficient of variation. SD: Standard deviation. n: number of replicates. CV under 10% was considered acceptable.

Detection of IgM, IgA and IgG antibodies against SARS-CoV-2 protein N was evaluated by analyzing 136 positive samples and 174 negative controls and ROC curve analyses were assessed to estimate the assay performance (Supplementary Figure 2A–C). The IgG direct-ELISA performed with sensitivity and specificity of 99.1% and 99.4%, whereas the IgA (56.6%, 99.4%) and the IgM (15.6%, 99.4%) showed much lower sensitivities. Significant correlations between the antibody response against RBD and protein N were observed (Supplementary Figure 2D–F).

**Figure 2.**
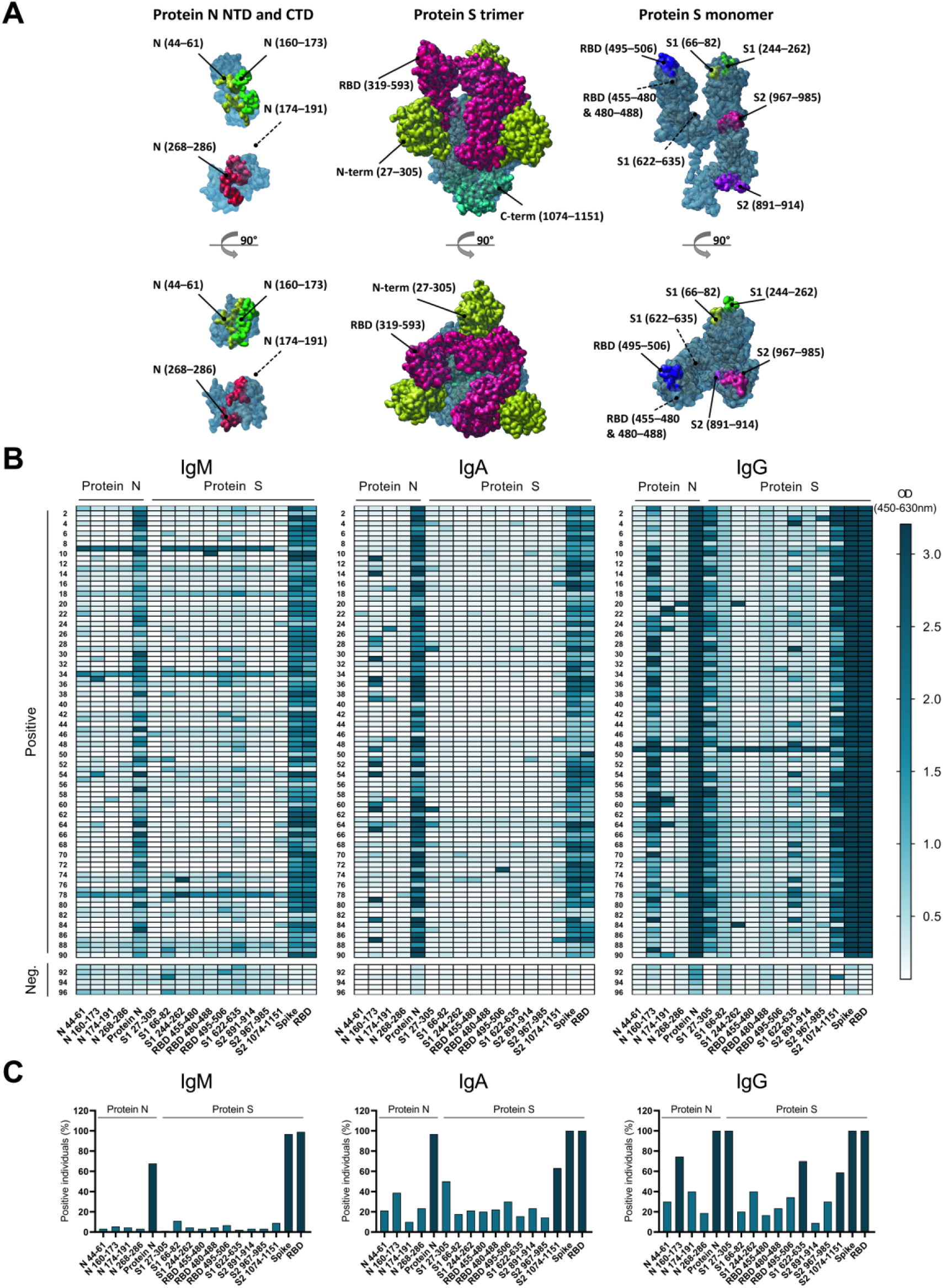
Differences in antibody seroconversion during SARS-CoV-2 infection and reactivity. (A) Location of the 14 protein fragments and short peptides on the protein S and protein N structures. The protein fragments are mapped on the atomic model of a partially open SARS-CoV-2 S trimer (6VSB) and the peptides in the monomer for clarity purposes. Protein N-derived short peptides are mapped on the freely oriented N-terminal RNA binding domain (6WKP) and the C-terminal dimerization domain (6WJI). Dashed lines represent approximate peptide locations not covered in the coordinate files. (B) Heatmap representation of OD of IgM, IgA and IgG antibodies binding to the 14 protein fragments and short peptides and protein S, protein N and RBD. (C) Simplified representation of positive bindings to the different proteins, protein fragments and short peptides. The cut off was stated by the mean of the negative controls plus three times the standard deviation. A total of 90 positive samples and 6 negative controls were analyzed.

### 3. Heatmap

To provide a better insight into antibody seroconversion during SARS-CoV-2 infection and reactivity against different locations on protein S and protein N, we conducted IgM, IgA and IgG detection in 90 positive samples against 14 protein fragments and short peptides located on the protein S and protein N structures, full-length RBD, protein S and protein N (Figure 2A). The heatmap shown in Figure 2B indicates a different reactivity of IgM, IgA and IgG towards the 17 proteins/peptides analyzed. Figure 2C represents a simplified overview of the % of individuals with antibodies recognizing each protein/fragment. A clear tendency towards a higher prevalence of IgG responses against the S1 subunit part of protein S and the middle part of protein N was observed. Parts of the S2 subunit and most of the N terminal part of RBD showed little immunogenicity. The cut off was calculated as the average of the negative controls plus three times the standard deviation.

### 4. Antibody detection in convalescent plasma

Levels of antibodies against SARS-CoV-2 were measured from plasma samples of 350 recovered individuals with COVID-19. The RBD S-ELISA measures total anti-RBD Igs present in the samples using a single dilution (Figure 3A), offering a sensitive and specific assay suitable for screening of the population. The direct-ELISAs offer a more detailed characterization of the antibody types and levels (Figure 3B–D) and the possibility to estimate antibody titers by interpolation. A total of 335 of the individuals previously infected with SARS-CoV-2 had mounted a detectable antibody response (Figure 3A), IgG being the most abundant isotype (present in the 330 individuals, Figure 3B). In comparison, responses of IgM and IgA isotypes were detected in 215 and 222 of the individuals, respectively (Figure 3C and Figure 3D).

**Figure 3.**
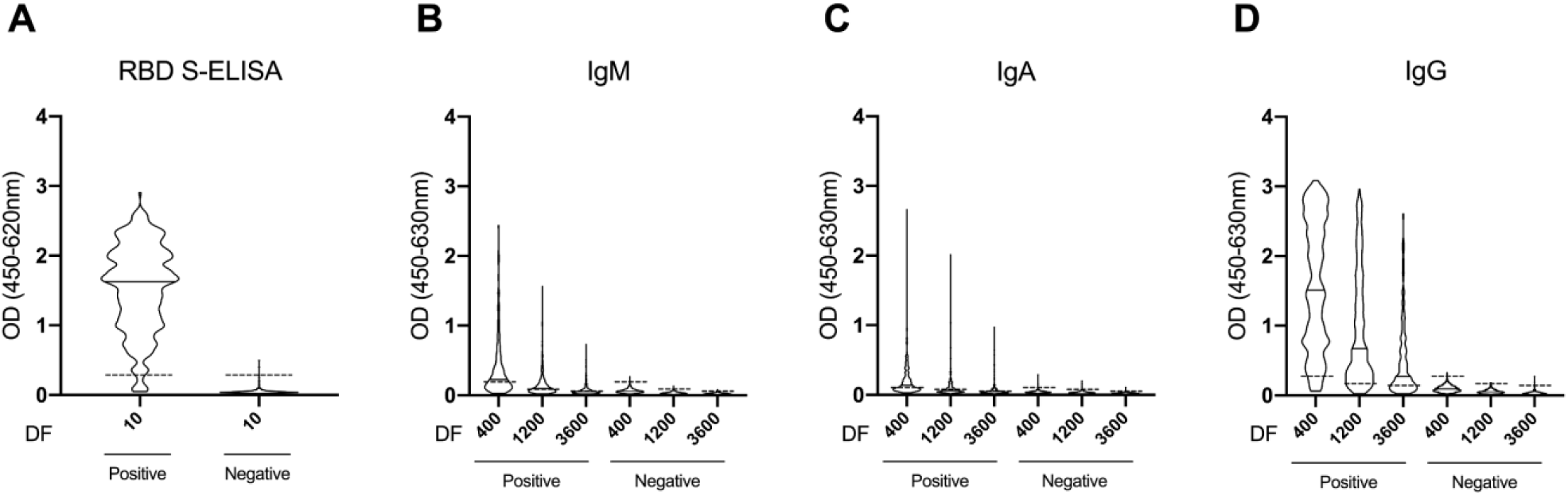
Antibody detection in convalescence plasma. OD levels of total Ig (A), IgM (B), IgA (C) and IgG (D) in convalescent individuals and negative controls. Samples were measured in a 1:10 dilution (A) or titrated thrice in a 1:400, 1:1200 and 1:3600 dilution (B-D). Dash line represents the cut off for each ELISA. A total of 350 positive samples and 580 negative controls were analyzed. DF: Dilution factor.

### 5. Antibody titers against SARS-CoV-2 are correlated with disease severity, symptom onset, age and symptomatology

A dynamic range of antibody titers expressed as arbitrary units (A.U.)/ml was obtained when OD values were interpolated by regression analysis using a four-parameter logistic curve fitting. The convalescent individuals were classified according to disease severity (Figure 4A–C) and symptom onset calculated as the time from the first self-perceived symptom related to COVID-19 to the moment of blood sampling (Figure 4D–F). We observed a highly significant difference in the antibody titers between the disease severity groups (Figure 4A–C) (*p* < 0.0001), with a clear association between increasing antibody levels and more severe disease symptomatology for all three antibody isotypes. IgG shows the most significant increase between the severity groups (Figure 4C). In contrast, asymptomatic individuals appeared not to follow the same severity tendency. This could be explained by the low number of individuals in this group, thus not being representative. When we assessed the difference in antibody titers between groups based on the time of symptom onset (Figure 4D–F), we observed a significant increase in IgG levels continuously over the time of sampling (Figure 4F), while IgM (Figure 4D) and IgA levels (Figure 4E) did not change significantly.

**Figure 4.**
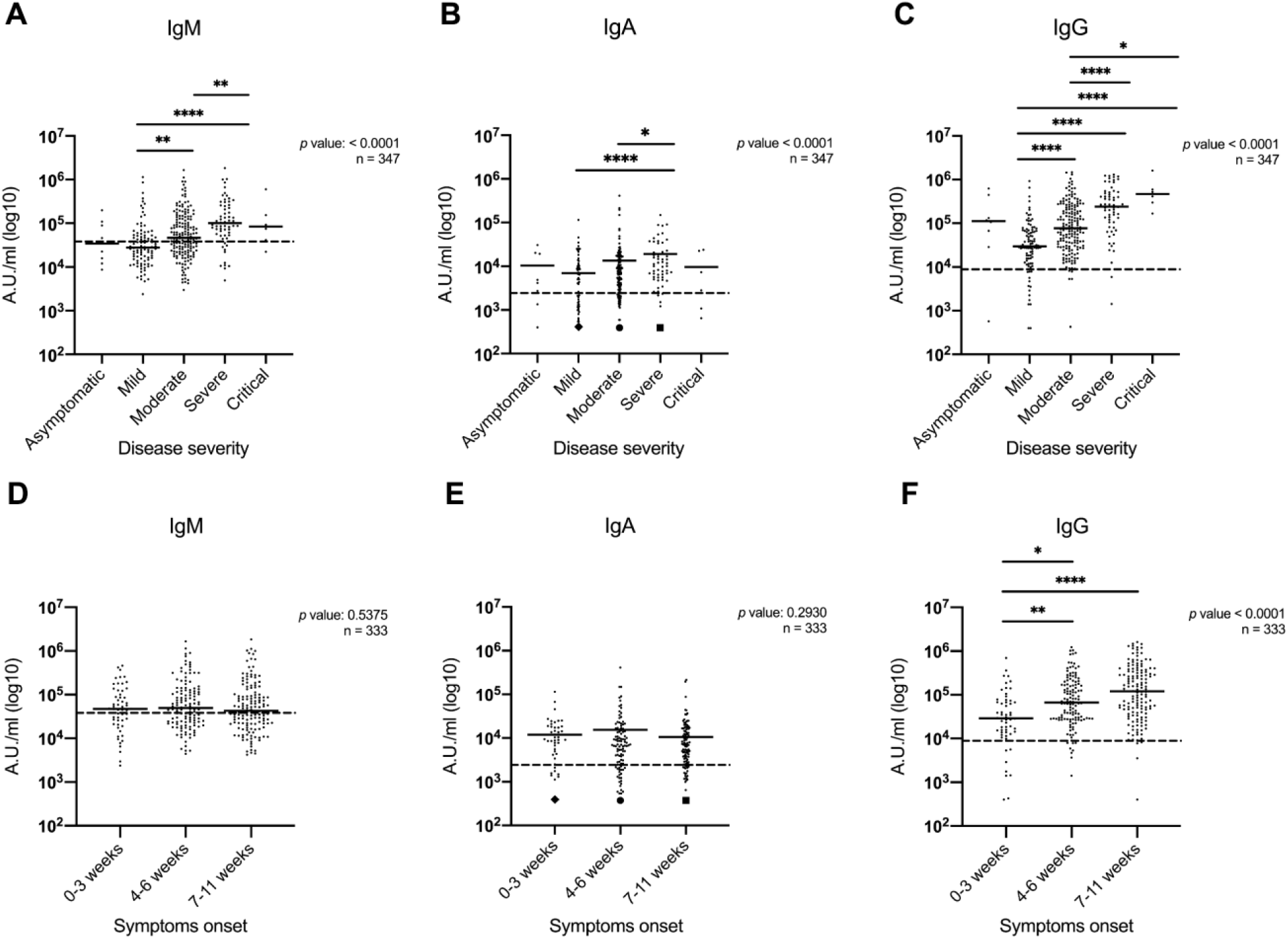
Antibody titers are correlated with disease severity and symptom onset. (A-C) Dynamic range represented in A.U./ml of IgM (A), IgA (B) and IgG (C) levels in convalescent individuals stratified accordingly to disease severity. (D-F) Dynamic range represented in A.U./ml of IgM (D), IgA (E) and IgG (F) levels in convalescent individuals stratified accordingly to disease severity. Dash line represents the cut off for each ELISA. In panel B, ♦ groups 35 individual samples, • groups 38 individual samples and ◼ groups 6 individual samples with A.U./ml value equal to 400. In panel E, ♦ groups 13 individual samples, • groups 24 individual samples and ◼ groups 40 individual samples with A.U./ml value equal to 400. Kruskal-Wallis test was performed. *p* value < 0.05 was considered significant. * = p < 0.05, ** = p < 0.01, *** = p < 0.001 **** = p < 0.0001.

Table 2 depicts the correlation between self-perceived COVID-19 symptomatology and the IgM, IgA and IgG titers, as well as the disease severity, sex and age. Symptoms such as fever, shortness of breath and lack of appetite were significantly and positively correlated with IgM and IgG levels. In contrast, IgA levels were significantly negatively correlated with the loss of sense of smell/taste and headache. Both age, sex (male) and disease severity were significantly positively correlated to the level of all three antibody isotypes. When adjusting the analysis between antibody levels and disease severity for age and sex, there was no longer a significant correlation between IgA and severity. Whereas it remained highly significantly correlated for IgM and IgG.

**Table 2.** Correlation between COVID-19 symptomatology and antibody levels of previously infected individuals with SARS-CoV-2

|  | IgM |  | IgA |  | IgG |  | Disease severity |  |
| --- | --- | --- | --- | --- | --- | --- | --- | --- |
|  | R<sup>2</sup> | p value | R<sup>2</sup> | p value | R<sup>2</sup> | p value | R<sup>2</sup> | p value |
| **No symptoms**<sup>a</sup> | -0.0334 | 0.5334 | -0.0034 | 0.9489 | -0.0056 | 0.9169 | -0.1011 | 0.0596 |
| **Fever**<sup>a</sup> | 0.1730 | 0.0012** | 0.0620 | 0.2483 | 0.2192 | <0.0001**** | 0.4281 | <0.0001**** |
| **Nasal congestion**<sup>a</sup> | -0.0995 | 0.0633 | -0.0534 | 0.3198 | -0.0845 | 0.1149 | -0.0523 | 0.3307 |
| **Pharyngalgia**<sup>a</sup> | -0.0935 | 0.0810 | -0.0764 | 0.1546 | -0.1236 | 0.0209* | -0.0427 | 0.4270 |
| **Coughing**<sup>a</sup> | 0.0227 | 0.6722 | 0.0139 | 0.7954 | 0.1013 | 0.0587 | 0.1382 | 0.0098** |
| **Shortness of breath**<sup>a</sup> | 0.1287 | 0.0161* | 0.0645 | 0.2293 | 0.2015 | 0.0002*** | 0.3508 | <0.0001**** |
| **Headache**<sup>a</sup> | -0.0128 | 0.8119 | -0.1099 | 0.0402* | -0.0869 | 0.1049 | 0.0477 | 0.3746 |
| **Muscle pain and joint pain**<sup>a</sup> | 0.0298 | 0.5785 | -0.0303 | 0.5727 | -0.0172 | 0.7491 | 0.1188 | 0.0266* |
| **Chest pain**<sup>a</sup> | -0.0383 | 0.4759 | -0.0008 | 0.9881 | 0.0324 | 0.5463 | 0.1356 | 0.0114* |
| **Fatigue**<sup>a</sup> | 0.1148 | 0.0320* | 0.0343 | 0.5233 | 0.0973 | 0.0695 | 0.2709 | <0.0001**** |
| **Lack of appetite**<sup>a</sup> | 0.2192 | <0.0001**** | 0.1723 | 0.0012** | 0.2644 | <0.0001**** | 0.3706 | <0.0001**** |
| **Nausea**<sup>a</sup> | -0.0101 | 0.8511 | 0.0632 | 0.2388 | 0.1389 | 0.0094** | 0.1821 | 0.0006*** |
| **Vomiting**<sup>a</sup> | -0.0122 | 0.8204 | -0.0238 | 0.6583 | 0.0622 | 0.2469 | 0.1309 | 0.0146* |
| **Diarrhea**<sup>a</sup> | -0.0040 | 0.9402 | 0.0556 | 0.3006 | 0.0819 | 0.1266 | 0.0815 | 0.1293 |
| **Abdominal pain**<sup>a</sup> | 0.0438 | 0.4142 | 0.0408 | 0.4470 | 0.0744 | 0.1654 | 0.1589 | 0.0029** |
| **Loss of sense of taste/smell**<sup>a</sup> | -0.0573 | 0.2853 | -0.1195 | 0.0256* | -0.0740 | 0.1677 | -0.0135 | 0.8026 |
| **Sex (M)**<sup>a</sup> | 0.1260 | 0.0185* | 0.1816 | 0.0007*** | 0.1239 | 0.0205* | 0.1403 | 0.0088** |
| **Age**<sup>a</sup> | 0.1972 | 0.0002*** | 0.3060 | <0.0001**** | 0.3618 | <0.0001**** | 0.2939 | <0.0001**** |
| **Disease severity**<sup>b</sup> | 0.3374 | <0.0001**** | 0.2662 | <0.0001**** | 0.4552 | <0.0001**** |  |  |
| **Disease severity**<sup>b,c</sup> | 0.147 | 0.006** | 0.053 | 0.330 | 0.310 | <0.0001**** |  |  |

Spearman rank correlation analysis was performed. *p* value < 0.05 was considered significant. \* = *p* < 0.05, \*\* = *p* < 0.01, \*\*\* = *p* < 0.001 \*\*\*\* = *p* < 0.0001.
<sup>a</sup> *n* = 349 participants.
<sup>b</sup> *n* = 348 participants.
<sup>c</sup> Partial correlation adjusted for age and sex.

## Discussion

We have developed an ELISA-based platform for detection SARS-CoV-2 antibodies comprising an indirect RBD S-ELISA for pan Ig detection and direct ELISAs for in-depth analyses of the IgM, IgA and IgG isotype responses towards RBD and protein N. RBD was chosen as the primary antigen for the screening and estimation of antibody titers for several reasons. It is regarded to be a sufficient representative part of protein S to induce an immunogenicity response (16) and we did not detect additional sensitivity improvements by employing the full protein S as a ligand-target in the direct ELISA (to be described in detail elsewhere). In a recent phase 1 trial, antibody responses against a vaccine candidate (S-2P antigen) and the RBD were assessed, finding similar Ig responses in pattern and magnitude between both antigens (17). One of the advantages of using RBD instead of full-length protein S is the more efficient production and higher stability of recombinant RBD due to its reduced size and simple tertiary structure. The relative unique primary sequence of the SARS-CoV-2 RBD (18) also reduces the risk of cross-reactive antibody signals derived from prior B cell responses against other types of coronaviruses. It is; however, important to annotate that this setup is highly flexible, making it possible to substitute the antigen upon a change in demand or in case of viral mutations. The use of protein N as a target antigen for serological screening has several theoretical pitfalls: the location of protein N makes it less accessible for B cell receptor interaction on the naïve B cell and probably requires a viral membrane degradation. Furthermore, protein N shows higher sequence conservation (90.5%) and thereby increase the risk of false-positive detection (19). Nevertheless, large commercial providers have chosen to use protein N as the serological target in different types of SARS-CoV-2 antibody assays (20).

In our study, the detection of IgM antibodies towards protein N or its fragments was weaker than IgG, suggesting a fast seroconversion of IgM into IgG. Furthermore, around 5–10% of the convalescent individuals did not mature any detectable antibody response, which is in good agreement with a previous study (21). The direct ELISA setup allows the use of different SARS-CoV-2 proteins in their full-length, shortened variants or fragmented immunogens, offering a useful tool to study the different reactivity patterns of IgM, IgA and IgG towards specific exposed areas on the viral antigens. We measured several different antigen areas on protein N and S to establish a heatmap of the antibody landscape. Based on the results, we could demonstrate a tendency towards immune dominating areas in the S1 unit and the central part of protein N. However, it is important to highlight that the heatmap does not represent the full sequences of the CoV antigens and that the dissection of antigens into shorter fragments could have a significant impact on the antibody reactivity. It could; however, in a more extensive setup, provide valuable information towards a targeted vaccine strategy to elucidate B cell epitopes relevant for the development of synthetically designed vaccine epitopes and select the best candidates based on the antibody titer upon clinical vaccine trials.

The indirect antigen RBD S-ELISA has applicatory value as a screening tool due to easy implementation, low reagent cost and a clear distinguished difference between positive and negative values. Serological surveys are the ideal tool for tracking the spread of an infectious disease within a population, particularly in the presence of asymptomatic cases or incomplete ascertainment of those with symptoms. The given specificity and sensitivity (99.7% and 94.9%, respectively) of the RBD S-ELISA indicate that the antibody test performed with good accuracy and can determine whether an individual has been infected with SARS-CoV-2. The direct ELISA setups, on the other hand, provide the best applications in a diagnostic setup for an in-depth examination of antibody-positive individuals. They allow quantification of IgM, IgA and IgG antibody levels, rendering more detailed information about the immunological response to the viral infection. The specificities of the direct assays were 99.1%, 99.3% and 99.5% for IgM, IgA, IgG, respectively. The assay sensitivities of the direct ELISAs are, especially for IgM and IgA, profoundly affected by the biological response, as a number of the convalescent individuals did not have measurable IgM levels or matured an IgA response. Nevertheless, when the three direct ELISAs were combined in a logistic regression analysis and ROC curve analysis the model predicts with 99.5% specificity and 94.3% sensitivity, which is very comparable with the RBD S-ELISA.

We show that IgG levels increases during the first 11 weeks after symptom onset, while the IgM and IgA levels remain stable for the same period of time. This was surprising as we would have expected a more pronounced decrease in IgM levels over time. This observation could indicate an importance of the two isotypes, reinforced by the fact that severity is correlated with high levels of antibodies of all three isotypes. It has previously been shown that both IgG independently and total antibody levels correlate with disease severity in patients during hospitalization (13), but to our knowledge, the prolonged clear correlation between IgA, IgM and IgG titers and disease severity have not been reported before. The data illustrates that individuals with mild symptoms during infection with SARS-CoV-2, in general, will mount a lower antibody response compared to individuals with moderate and severe symptoms several months after recovering from a COVID-19 infection. This observation gives an essential insight into the immunological response regarding clinical disease presentation, which further highlights the demand for more quantitative assays.

In this study, all three antibody isotypes correlate positively with age and sex (male), which can be explained by the fact that severe COVID-19 infection is more pronounced in the elderly male population (22). We show that the antibody titers, especially IgG levels, are correlated with specific symptom characteristics, including fever, pharyngalgia, shortness of breath and nausea, which again shows the link between clinical manifestations and the immunological response. The IgA level, on the other hand, was negatively correlated with loss of sense of taste/smell and surprisingly showed no other correlation to the symptomatology of the upper respiratory tract in this study. The role of IgA, which is considered the predominant antibody involved in mucosal immunity, remains to be fully understood. IgA is suggested to mediate anti-viral defense functions at different anatomic levels in relation to mucosal epithelium (23). However, the mechanisms behind this remain unknown and often gain limited attention during infectious studies. In this respect, it could be interesting to examine the IgA levels in mucosal tissue during SARS-CoV-2 infection and determine whether the mucosa-associated IgA plays a significant role during SARS-CoV-2 infection.

Our findings provide support to the notion that antibodies towards SARS-CoV-2 represent a double-edged sword. Antibodies are important in viral neutralization, but also in Fc receptor-mediated phagocytosis, antibody-dependent cellular cytotoxicity (ADCC) and complement-dependent cellular cytotoxicity (CDCC) and subsequent elimination of pathogens. However, it is known that particularly ADCC and CDCC can drive harmful and systemic pro-inflammatory responses that can have severe pathophysiological consequences. Thus, based on our findings and others, it may be suggested that an unwanted immune response towards SARS-CoV-2 may be one of the mechanisms causing hyperactivation of macrophages and monocytes, leading to the deadly cytokine storm, which seems to be a hallmark of COVID-19 (24).

Whether a previously infected individual can expect stable long-term protection against reinfection with SARS-CoV-2 remains unknown. A recent study found a correlation between the production of neutralizing antibodies against RBD and elevated IgG titers in convalescence COVID-19 individuals (25), reinforcing the use of RBD as the candidate to analyze for neutralizing antibodies in these individuals. The durability of neutralizing antibodies (primarily IgG) against SARS-CoV-2 has yet to be defined, but persistence for up to 40 days from symptom onset has been described (13, 26). In comparison, when following infection with SARS-CoV-1, concentrations of IgG remained high for approximately 4 to 5 months before subsequently declining slowly during the next 2 to 3 years (27). It is uncertain whether an individual with low antibodies titers, mainly IgG, has a higher risk of reinfection compared to an individual with high levels of antibodies post disease. It has recently been suggested that in the absence of antibody levels, an individual could be protected against reinfections by the presence of memory T cells (28). Furthermore, the major proportion of convalescent individuals, included in this study, indeed show a dominating IgG response, suggesting that both affinity maturation, isotype class switching and B cell memory response has occurred. These B cell populations could, together with the memory CD4+ and CD8+ cells, secure a fast and efficient response to a secondary exposure of SARS-CoV-2.

This study is limited by relying on participants’ retrospectively self-reported symptomatology and symptom debut, which allows for an unknown amount of misclassification. Moreover, with this design, we could not monitor the antibody response concerning survival. However, because of the detailed analysis of the antibody responses, and the clear associations despite the retrospective design, we assume that the associations would be even stronger in a carefully conducted prospective designed study.

In conclusion, we have established robust, flexible and specific ELISA-based platforms for detection SARS-CoV-2 antibodies and presented novel insight into the link between antibody responses and COVID-19 disease severity.

## Methods

### 1. Buffers

The following buffers were used: PBS (10.1 mM Na_2_HPO_4_, 1.5 mM KH_2_PO_4_, 137 mM NaCl, 2.7 mM KCl), PBS-Tween (PBS-T) [PBS, 0.05% T-20 (8221840050, Merck, Darmstadt, Germany)] and PBS-T-EDTA + skim milk (dilution buffer) [PBS, 0.05% T-20, 5 mM EDTA (EDS-500G, Merck), 5% Skim milk (70166, Merck)].

### 2. SARS-CoV-2 antigens

The coding sequence for protein S RBD (QIC53204.1, aa R319–S593) was synthesized by GenScript and cloned into a pcDNA3.4 expression vector with an N-terminal human VH1-2 signal peptide and a C-terminal 10xHis tag followed by an AviTag (-GSG-HHHHHHHHHH-GSG-GLNDIFEAQKIEWHE). The sequence of protein N (QLD29192.1) was optimized as described elsewhere (29) and synthesized by GeneArt (Thermo Fisher Scientific, Waltham, MA, USA) into a pcDNA3.4 vector with the human serum albumin signal peptide and a C-terminal 8xHis tag (-GS-HHHHHHHH). Both constructs were expressed using a mammalian transient expression system. On day five after transfection, the supernatants were harvested by centrifugation and sterile filtered. The recombinant proteins were purified using a 2-step automated purification method setup on an ÄKTA Express chromatography system with an immobilized metal affinity Excel HisTrap column and a size exclusion Superdex200 column (chromatography system and columns Cytiva, Marlborough, MA, USA). The purified proteins were stored in a buffer composed of 20 mM Hepes, 150 mM NaCl, pH 7.4. A portion of the RBD was specifically biotinylated in the AviTag sequence using a BirA500 kit (Avidity LLC, CO, USA).

### 3. Convalescent individuals samples and healthy individuals’ controls

A total of 350 recovered individuals previously tested RT-PCR positive for SARS-CoV-2 were included in the study. The Department of Cardiology at Herlev University Hospital in Denmark recruited the participants. The RT-PCR positive participants are comprised of males and females aged from 18–86 and course of disease ranged from asymptomatic to critically ill. All participants were invited to complete an electronic self-report questionnaire providing additional information about symptom onset, characteristics and disease severity divided into the following groups: asymptomatic, mild, moderate, severe or critical. The mild disease was defined as having few symptoms and generally feeling well, moderate disease as being bedridden at home, severe disease as the need for hospitalization and critical disease as need of admission to the intensive care unit (ICU) for mechanical ventilation. Characteristics of the study participants are detailed in Table 3. Serum and EDTA plasma samples were stored in aliquots and kept frozen at −80 °C until used. A total of 580 EDTA plasma samples collected from healthy blood donors at four-time points (May 2017, February 2018, September 2018 and February 2019) were used as negative controls for SARS-CoV-2. All SARS-CoV-2-positive samples and controls, a total of 930 samples, were subjected to antibody detection in the different assays as described below.

**Table 3.** Characteristics of the cohort of previously infected individuals with SARS-CoV-2

| Variables | Entire cohort (n = 350) |
| --- | --- |
| Age, years | 52 (41–63) |
| Sex, female (%) | 199 (56.9) |
| **Disease severity<sup>a</sup>** |  |
| Asymptomatic (%) | 9 (2.6) |
| Mild (%) | 96 (27.4) |
| Moderate (%) | 172 (49.1) |
| Severe (%) | 62 (17.7) |
| Critical (%) | 6 (1.7) |
| **Symptom characteristics<sup>b</sup>** |  |
| No symptoms (%) | 8 (2.3) |
| Fever (%) | 244 (69.9) |
| Nasal congestion (%) | 130 (37.2) |
| Pharyngalgia (%) | 143 (41) |
| Coughing (%) | 227 (65) |
| Shortness of breath (%) | 136 (39) |
| Headache (%) | 231 (66.2) |
| Chest pain (%) | 201 (57.6) |
| Muscle and joint pain (%) | 88 (25.2) |
| Fatigue (%) | 274 (78.5) |
| Loss of appetite (%) | 172 (49.3) |
| Nausea (%) | 86 (24.6) |
| Vomiting (%) | 26 (7.4) |
| Diarrhea (%) | 98 (28.1) |
| Abdominal pain (%) | 48 (13.8) |
| Loss of sense of taste/smell (%) | 200 (57.3) |

Values denote median (interquartile range) or number (%).
<sup>a</sup> Participants with missing values n = 5.
<sup>b</sup> Participants with missing values n = 1.

### 4. ELISA-based assays

#### 4.1. RBD sandwich ELISA: In-house produced assay measuring a total Ig antibody response against SARS-CoV-2

Nunc™ MaxiSorp Flat-Bottom 96-Well plates (442404, Thermo Fisher Scientific) or Clear Flat-Bottom Immuno Nonsterile 384-Well Plates (464718, Thermo Fisher Scientific) were coated with 0.5 µg/ml RBD in PBS overnight (ON) at 4°C. Within the 384-well setup, plates were blocked with WellChampion blocking solution (4900, KemEnTec Diagnostics, Taastrup, Denmark) following the manufacturer’s instructions. Within the 96-well setup, plates were blocked with PBS-T for 1 h. Samples were diluted 1:10 (384-wells) or 1:100 (96-wells) in PBS-T, applied to the plates and incubated for 1.5 h at RT. Antibodies anti-SARS-CoV-2 were detected using Pierce^™^ HRP-conjugated streptavidin (21130, Thermo Fisher Scientific) diluted 1:16000 in PBS-T was mixed with 0.5 µg/ml biotin-labelled RBD and incubated for 2 h RT. TMB ONE (4380A, KemEnTec Diagnostics) was used as a substrate and allowed to react for 5 minutes. The reaction was stopped with 0.3 M H_2_SO_4_ and the OD of the samples was measured at 450-620 nm using a Tecan Magellan reader for 96-well plates (Tecan, Männedorf, Switzerland) and Multiscan FC reader for 384-well plates (Thermo Fisher Scientific). Plates were washed four times with PBS-T between the steps mentioned above. All 384-well plates were handled by the Tecan Fluent automated workstation.

#### 4.2. Direct ELISA: In-house produced assay measuring SARS-CoV-2 IgM, IgA and IgG antibody levels

Nunc™ MaxiSorp Flat-Bottom 96-Well plates or Nunc™ MaxiSorp Flat-Bottom plates nonsterile 384-well plates were coated with 1 µg/ml RBD or protein N in PBS ON at 4°C. Positive and negative samples were diluted 1:400 in dilution buffer, diluted in a 3-fold to 1:1200 and 1:3600 and incubated 1 h shaking at RT. EDTA plasma from a recovered COVID-19 patient with high titers of IgM, IgA and IgG used as a calibrator was applied in a 2-fold serial dilution and incubated as stated above. Antibodies bound to SARS-CoV-2 antigens were detected using 0.5 µg/ml HRP-conjugated polyclonal rabbit antibodies against human, human IgM (P0215), human IgA (P0216) or IgG (P0214) (all from Agilent Technologies, Santa Clara, CA, USA) diluted in PBS-T and incubated for 1 h shaking at RT. TMB ONE substrate was applied and allowed to react for 10 minutes for IgM and IgA and 7 minutes for IgG. The reaction was stopped with 0.2 M H_2_SO_4_ and the OD was measured at 450-630 nm using a Synergy HT Absorbance Reader (Bio-Tek, Winooski, VT, USA). Plates were washed four times with PBS-T between the steps mentioned above. All 384-well plates were handled by the Biomek FX Automated Workstation (Beckman Coulter, Brea, CA, USA).

### 5. Assay development and validation: Specificity of the signal, limit of detection, variation, parallelism and clinical performance

The RBD S-ELISA and the direct ELISAs was subjected to optimization with regards to dilution range, the use of blocking buffers and variations in incubation times. The final conditions were chosen based on S/N ratios. Specificity and sensitivity were calculated based on ROC curve representation and selection of the most appropriate cut off by prioritizing the specificity. Assay sensitivity regarding the limit of detection was determined by the concentration given by interpolating the OD value of the cut off. The calibrator was prepared by spiking 100 µg/ml of recombinant human monoclonal IgG antibody against SARS-CoV-2 Spike1 (A02038, GenScript, Piscataway, NJ, USA) into normal human serum and diluting in serum into a 2-fold dilution. Samples were treated as patient samples and further diluted 1:100 in the S-ELISA and 1:1000 in the direct ELISA in PBS-T followed by incubated as stated above. Intra-assay variation was evaluated by calculating the coefficient of variation (CV) of an individual CVs for all the duplicates in a total of 40 samples. Inter-assay variation was evaluated by calculating the CV of two samples run in duplicates in 8 separate plates on three different days. The parallelism between serum and plasma samples was evaluated by comparing 137 pairs of serum and plasma samples using Spearman rank correlation tests. To evaluate whether the antibody levels correlated with the disease severity and/or the days after symptom onset, sample absorbances were logistically transformed and a four-parameter logistic curve fitting was applied to calibrate the antibody levels into A.U./ml. The appropriate dilution for each sample was chosen based on the best fit in the linear range of the calibrator. The interpolated value in A.U./ml was corrected by the dilution factor (400, 1200 or 3600). A sample OD value below 0.06 was automatically given the value of 1 A.U./ml.

### 6. Heatmap

A total of 12 different SARS-CoV-2 protein fragments on the protein S1 and S2 and protein N coupled via an N-terminal cysteine and maleimide conjugation to recombinant-human serum albumin (rHSA) (AlbIX, Novozymes, Bagsværd, Denmark), 2 short proteins from protein S and full-length protein S, protein N and RBD were analyzed for immunogenicity capacity on the direct ELISA. Proteins details are illustrated in Figure 2A. Nunc™ MaxiSorp Flat-Bottom plates nonsterile 384-Well Plates were coated with 1 µg/ml of the proteins in PBS ON at 4°C. A total of 90 RT-PCR positive samples and six negative controls were diluted 1:20 in dilution buffer and incubated as mention above. Detection and development procedures were followed, as described in subsection 4.2.

### 7. Statistics

Statistical analyses were performed using GraphPad Prism version 8.0 (GraphPad Software, La Jolla, CA, USA) and Statistical Package for the Social Sciences 25.0 software (SPSS Inc., Chicago, IL, USA). Measurements of samples and the calibrator were performed in duplicates. Data were analyzed for normal distribution using the Anderson-Darling test. Estimation of levels of IgM, IgA and IgG were interpolated by regression analysis using a four-parameter logistic curve fitting and results were given in A.U./ml (in a 1:200 dilution, the calibrator was defined to contain 2000 A.U./ml). Statistical differences between disease severity and symptoms onset groups were analyzed using one-way ANOVA (Kruskal-Wallis test) with Dunn’s multiple comparison test. Spearman rank correlation tests were used to determine the correlation between different experimental parameters. Data are represented as the average of sample duplicate and the median. Significance levels: * = p < 0.05, ** = p < 0.01, *** = p < 0.001 **** = p < 0.0001 and p < 0.05 was considered statistically significant.

### 8. Study approval

All procedures involving the handling of human samples are in accordance with the principles described in the Declaration of Helsinki and ethically approved by the Regional Ethical Committee of the Capital Region of Denmark (H-20028627).

## Data Availability

The article consists solely of original data and we have all raw files to prove it.

## List of abbreviations

ACE2: Angiotensin-converting enzyme 2 receptor
A.U.: Arbitrary units
CoV: Coronavirus
COVID-19: Coronavirus Disease 2019
CV: Coefficient of variation
Ig: Immunoglobulin
MERS: Middle East respiratory syndrome
OD: Optical density
ON: Overnight
Protein N: Nucleocapsid protein
Protein S: Spike protein
RBD: Receptor binding domain
rHSA: Recombinant human serum albumin
ROC: Receiver operating characteristic
RT: Room temperature
S/N: Signal to noise
SARS: Severe acute respiratory syndrome
S-ELISA: Sandwich ELISA

## Author contributions

CB.H, I.J and L.PA developed the direct setup and analyzed the data. LH.L and M.L performed the RBD S-ELISA. JG.J, C.H, JR.B and T.E enabled recombinant antigen production. CS.J provided essential information for assay validation. K.I. provide convalescence plasma from previously infected individuals with SARS-CoV-2. R.BO, MO.S and P.G designed the study. CB.H, I.J, L.PA, A.R, R.BO, P.G and MO.S wrote the manuscript. All authors critically reviewed the manuscript.

## Acknowledgements

The authors thank Jytte Bryde Clausen and Bettina Eide Holm for excellent technical assistance and José Juan Almagro Armenteros (University of Copenhagen, Copenhagen, Denmark) for his statistical advice. This work was financially supported by the Carlsberg Foundation (CF20-0045) and the Novo Nordisk Foundation (205A0063505).

